# Demography, social contact patterns and the COVID-19 burden in different settings of Ethiopia: a modeling study

**DOI:** 10.1101/2020.11.24.20237560

**Authors:** Filippo Trentini, Giorgio Guzzetta, Margherita Galli, Agnese Zardini, Fabio Manenti, Giovanni Putoto, Valentina Marziano, Worku Nigussa Gamshie, Ademe Tsegaye, Alessandro Greblo, Alessia Melegaro, Marco Ajelli, Stefano Merler, Piero Poletti

## Abstract

**Background:** COVID-19 spread may have a dramatic impact in countries with vulnerable economies and limited availability of, and access to, healthcare resources and infrastructures. However, in sub-Saharan Africa a low prevalence and mortality have been observed so far.

**Methods:** We collected data on individuals’ social contacts in Ethiopia across geographical contexts characterized by heterogeneous population density, work and travel opportunities, and access to primary care. We assessed how socio-demographic factors and observed mixing patterns can influence the COVID-19 disease burden, by simulating SARS-CoV-2 transmission in remote settlements, rural villages, and urban neighborhoods, under the current school closure mandate.

**Results:** From national surveillance data, we estimated a net reproduction number of 1.62 (95%CI 1.55-1.70). We found that, at the end of an epidemic mitigated by school closure alone, 10-15% of the overall population would have been symptomatic and 0.3-0.4% of the population would require mechanical ventilation and/or possibly result in a fatal outcome. Higher infection attack rates are expected in more urbanized areas, but the highest incidence of critical disease is expected in remote subsistence farming settlements.

**Conclusions:** The relatively low burden of COVID-19 in Ethiopia can be explained by the estimated mixing patterns, underlying demography and the enacted school closures. Socio-demographic factors can also determine marked heterogeneities across different geographical contexts within the same country. Our findings can contribute to understand why sub-Saharan Africa is experiencing a relatively lower attack rate of severe cases compared to high income countries.

## Introduction

Despite limited access to healthcare^1,2^ and relatively milder social distancing restrictions compared to those imposed in most high-income countries^3,4^, COVID-19 mortality rates have been relatively low throughout Africa^5^. As of November 5, 2020, the World Health Organization (WHO) reports 44,012 deaths out of 1,838,867 diagnosed cases in the continent, for a case-fatality ratio (CFR) of 2.4%, compared to about 3% in the rest of the world^5^. However, SARS-CoV-2 transmission dynamics have been highly heterogeneous across different African countries in terms of timing and implemented interventions^6^.

In sub-Saharan Africa, Ethiopia is second only to South Africa in terms of number of recorded cases and deaths, with an overall CFR of about 1.5%^5^. The first COVID-19 case was confirmed on March 13, 2020 and, less than a month later, the Ethiopian Prime Minister declared a state of emergency in the country on April 8, 2020^7^. Since then, borders and school closure were implemented, public institutions and firms operated at minimum capacity or under complete closure, and people were advised to stay at home^8^. Despite these restrictions, several hundred cases have been reported in all the 12 regions of Ethiopia^9^. The possible spread of SARS-CoV-2 in rural areas of the country is especially dangerous because of the sparse presence of well-resourced health facilities implying long travel distances for remote populations, which is an important barrier to universal access to primary care^2^. Moreover, the healthcare workforce in Ethiopia is 5 times lower than the minimum threshold defined by the WHO for Sustainable Development Goals health targets^10^, and far below the African average^11^.

Recent modeling studies investigated the impact of control measures, such as self-isolation and temporary lockdowns, in a number of sub-Saharan African countries, highlighting the difficulties in defining effective, feasible and sustainable strategies for suppression or mitigation of COVID-19 epidemics^12,13,14,15^. In this work, we aim to assess how demographic factors and age-specific mixing patterns can influence COVID-19 burden in Ethiopia across different geographical contexts characterized by different levels of access to healthcare.

## Methods

### Study design

Between November and December 2019, we conducted a survey based on individual interviews to estimate age-specific mixing patterns in four districts (*woreda*) of the South West Shewa Zone (SWSZ) of the Oromia Region, Ethiopia. About 40% of the SWSZ population is below 15 years of age and about 68% lives in remote rural settlements, 18% in rural villages, and 14% in the largest town of the area (Woliso Town, 53,065 inhabitants). The districts targeted by our study encompass a population of 449,460 inhabitants and represent the main catchment area of the St. Luke Hospital located in Woliso Town, a well-resourced health facility acting as the referral hospital for the entire Zone^2^.

The study consists in a cross-sectional survey with two-stage stratified random sampling by location and age group. The survey was conducted in eight different sites, choosing two neighborhoods (*kebele*) for each district under study, in such a way to capture contact patterns in areas characterized by different population density, work and travel opportunities, and access to the healthcare infrastructure. Three types of geographical contexts were considered: remote settlements (consisting of scattered subsistence farming settlements), rural villages (consisting of concentrated clusters of households served by a main road, and better access to main public services), and urban neighborhoods inside Woliso Town (significantly higher population density and full access to public services^16^).

For each site, a target sample size of 105 study participants was set on the basis of findings from previous contact surveys^17,18^ to provide the desired precision in the mean number of contacts (see Appendix S1). Households and study participants were randomly sampled using predefined quotas for each site, sex, and age group. A household was defined as a group of individuals living under the same roof and sharing the same kitchen on a daily basis. One individual per household was interviewed. If the study participant was a student, additional shorter interviews were performed to complement the data with information about close contacts occurring at school.

### Data collection

Participants were asked to recall information on the frequency, location, type of social encounters from the day preceding their interview, providing the age (or age range when exact age was unknown) and their relationship for each listed contact. A contact was defined as an interaction between two individuals, either physical (when involving skin-to-skin contact), or non-physical (when involving a two-way conversation with five or more words in the physical presence of another person, but no skin-to-skin contact)^17,18^. The participants’ age, sex, education and occupational status were recorded along with details on their household composition. In the SWSZ, schools may host up to 100 students within a single class. To avoid inaccurate reporting of the number of school contacts, participants were only asked to count the total number of physical contacts they had at school in the previous day, without further details. Information on the age of students attending the targeted schools for different grades was also collected. Schools were regularly open during the survey period.

### Contact patterns and data analysis

For each type of geographical context, we computed the mean number of contacts reported by respondents after grouping by age (six 10-year age groups from 0 to 59 years and one age group for individuals aged 60 years or older) and by contact setting (households, schools, and the general community). Since for many study participants it was difficult to distinguish encounters occurred because of their job from other random contacts, all social interactions occurring outside family and schools were aggregated with contacts occurring in the general community. Age-specific contact matrices were computed considering both physical and non-physical contacts. Variability due to sampling of study participants was explored by computing 1,000 bootstrapped contact matrices, where each bootstrap consisted in sampling with replacement a number of interviews equal to the original sample size, choosing the age of the participant with probability proportional to the Ethiopian age distribution^19^. The proportions of the SWSZ population living in remote settlements, rural villages and in urban neighborhoods were used as sampling weights to compute an average contact matrix for the entire SWSZ. Full details about the study design, data collection and the analysis of contact patterns are provided in the Appendix S1.

### Transmission model

We simulated SARS-CoV-2 spread in the different geographical contexts of the SWSZ, using an age-structured SIR compartmental model with three consecutive stages of infectiousness, in such a way to reproduce a gamma-distributed generation time of mean 6.6 days^20^. The model was run separately for each geographical context, using estimates of the population age structure and of the age-specific contact matrix computed from survey data (see Appendix). Because school closure in all of Ethiopia was mandated much before the exponential growth of reported COVID-19 cases, we included only data on household and community contacts in the contact matrices. However, we included school contacts to estimate the theoretical SARS-CoV-2 transmission potential in the absence of a school closure mandate. We considered susceptibility to SARS-CoV-2 infection to vary with age. We adopted posterior distributions for the relative probability of developing infection upon effective exposure to an infectious case as previously estimated in Zhang et al.^19^, where the age-group 15-64 years is taken as a reference; an average relative susceptibility of 0.33 (95%CI: 0.24-0.47) was estimated for children under 15 years of age, and of 1.47 (95%CI: 1.16-2.06) for older adults (above 65 years)^19^. These estimates are aligned with other independent studies (reviewed in Viner et al.^21^). We assumed the same infectiousness across individuals of different ages. Two separate sensitivity analyses were carried out assuming 1) homogeneous susceptibility by age; 2) a lower infectiousness of children (see Appendix).

We estimated the SARS-CoV-2 net reproduction number from the curve of reported cases in Ethiopia during the phase of exponential growth^5,22^ (see Appendix). We then set a per-contact transmission rate in the model such that the reproduction number, calculated using the next-generation matrix approach^23^ matches the obtained national estimate (see Appendix). A sensitivity analysis was conducted to explore the effect of a 20% increase or a 20% decrease of the reproduction number on COVID-19 burden.

We computed projections of the number of SARS-CoV-2 infections, cases with respiratory symptoms or fever, and COVID-19 critical cases (either requiring mechanical ventilation or resulting in a fatal outcome), based on available estimates of the age-specific risks^24^.

## Results

### Social contact data

A total of 938 study participants were interviewed with 43% of them living in rural remote settlements, 35% in rural villages, and 22% from urban neighborhoods (Table 1). 227 participants were students, 22.9% of whom were between 5 and 9 years of age, 71.8% between 10 and 19 years, and 4.9% older. School attendance rates among the study participants aged 5-18 years was 67%, 80% and 77% in remote, rural and urban sites, respectively. The median class size ranged from 70 children per class in rural villages to 90 in remote settlements. Only 27% of our study participants reported travels outside their village in the last month; 87.3% reported they were never admitted to the local hospital (see Appendix S1).

**Table 1.** Characteristics of study participants.

**Number of study participants**
| **VARIABLE\*** | **OVERALL**<br>**n (%)** | **REMOTE**<br>**n (%)** | **RURAL**<br>**n (%)** | **URBAN**<br>**n (%)** |
| --- | --- | --- | --- | --- |
| **Total** |  |  |  |  |
|  | 938 (100.0) | 400 (42.6) | 326 (34.8) | 212 (22.6) |
| **Age** |  |  |  |  |
| <10y | 382 (40.7) | 160 (40) | 137 (42) | 85 (40.1) |
| 10-19y | 198 (21.1) | 85 (21.2) | 66 (20.2) | 47 (22.2) |
| 20-29y | 92 (9.8) | 40 (10) | 32 (9.8) | 20 (9.4) |
| 30-39y | 117 (12.5) | 50 (12.5) | 42 (12.9) | 25 (11.8) |
| 40-49y | 59 (6.3) | 26 (6.5) | 18 (5.5) | 15 (7.1) |
| 50-59y | 40 (4.3) | 17 (4.2) | 13 (4) | 10 (4.7) |
| 60y + | 50 (5.3) | 22 (5.5) | 18 (5.5) | 10 (4.7) |
| **Occupation** |  |  |  |  |
| Student | 226 (24.1) | 85 (21.2) | 87 (26.7) | 54 (25.5) |
| Office/Shop worker | 47 (5.0) | 4 (1.0) | 25 (7.7) | 18 (8.5) |
| Agriculture | 112 (11.9) | 84 (21) | 25 (7.7) | 3 (1.4) |
| Unemployed/Retired | 44 (4.7) | 9 (2.3) | 12 (3.7) | 23 (10.8) |
| Other | 509 (54.3) | 218 (54.5) | 177 (54.3) | 114 (53.8) |
| **Sex** |  |  |  |  |
| Female | 478 (51) | 206 (51.5) | 170 (52.1) | 102 (48.1) |
| Male | 460 (49) | 194 (48.5) | 156 (47.9) | 110 (51.9) |

\* No missing data for any of the three listed variables.

Age and sex were also recorded for all the 4,635 household members of the 938 study participants. The mean household size in remote settlements was 5.5 (95% CI: 5.3-5.7), significantly larger (Tukey test p<0.001) than in rural villages (4.6, 95% CI: 4.4-4.8) and in urban neighborhoods (4.4, 95%CI: 4.2-4.6), while no significant difference in the household size was found between the latter two settings (Tukey test p=0.48).

Overall, 5,690 non-school contacts were reported by the 938 study participants (median 6 contacts per person, range 1-26, see Table 2). Of these, 79.9% were physical and 43.0% involved a single social interaction during the day.

**Table 2.**
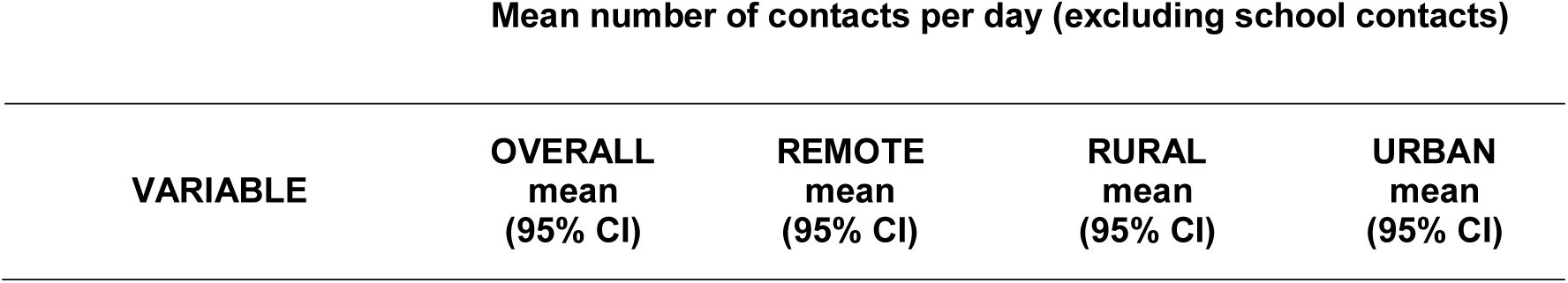
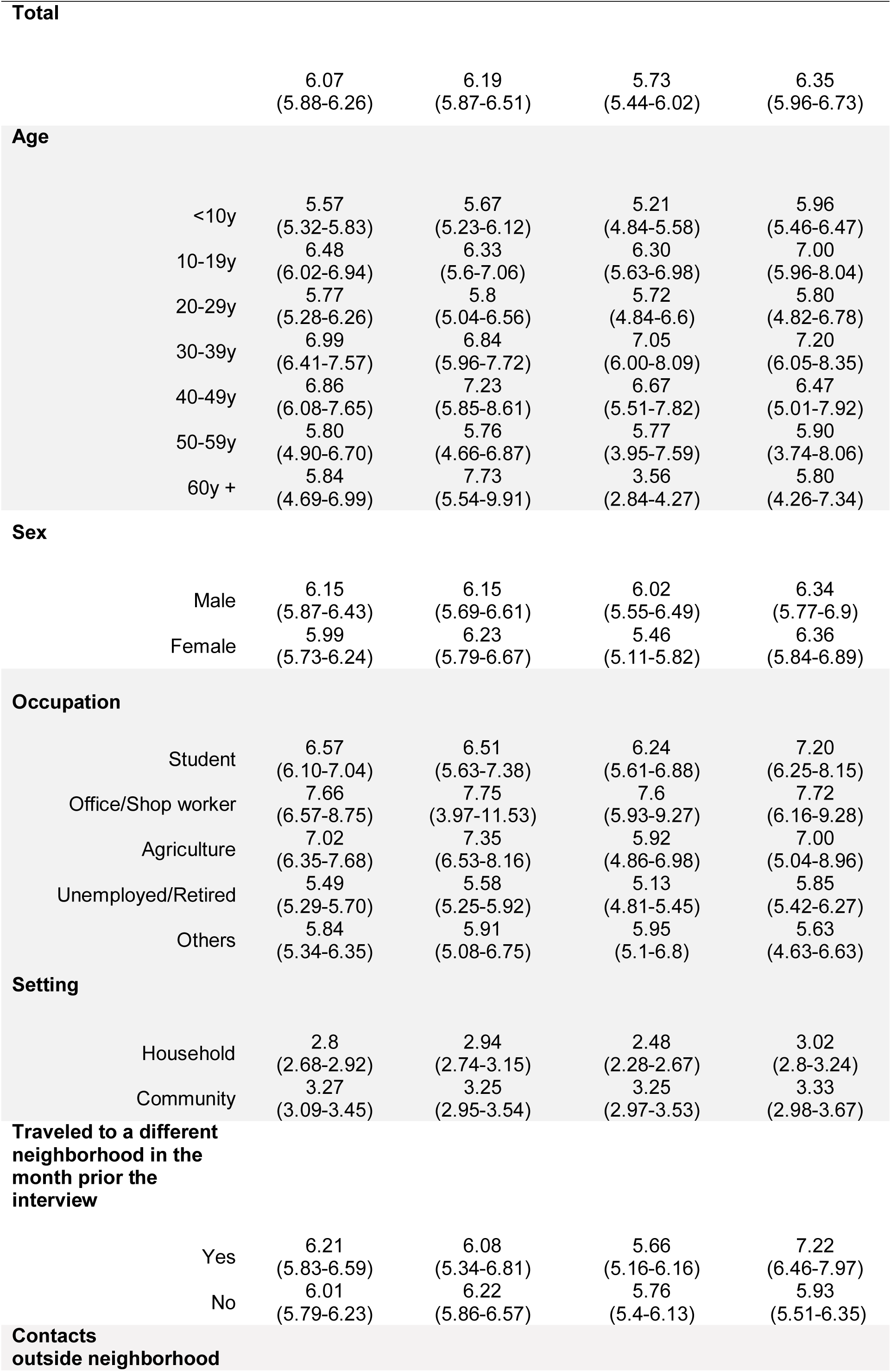
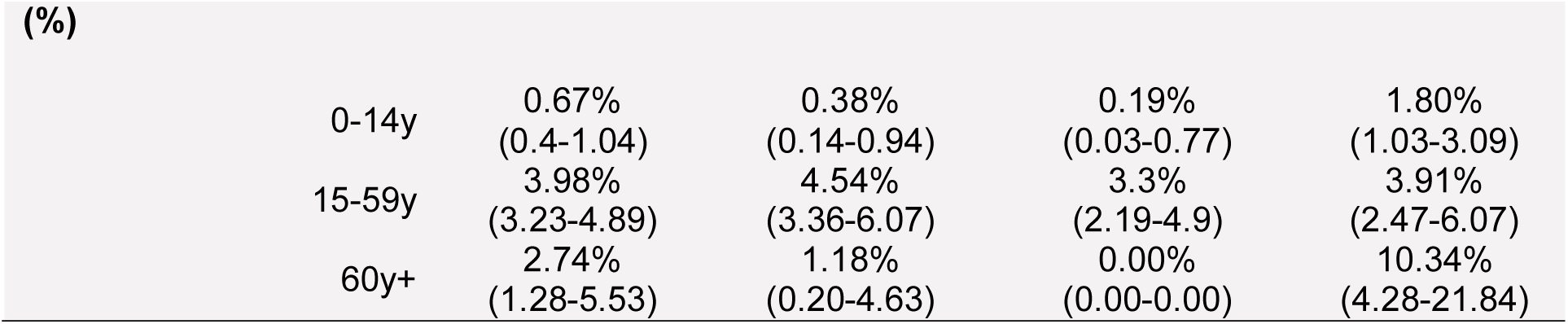
Mean number of recorded daily contacts, excluding contacts at school, by age, across different geographical contexts.

| **Mean number of contacts per day (excluding school contacts)** |  |  |  |  |
| --- | --- | --- | --- | --- |
| **VARIABLE** | **OVERALL**<br>**mean**<br>**(95% CI)** | **REMOTE**<br>**mean**<br>**(95% CI)** | **RURAL**<br>**mean**<br>**(95% CI)** | **URBAN**<br>**mean**<br>**(95% CI)** |

|  |  |  |  |  |  |
| --- | --- | --- | --- | --- | --- |
| **Total** |  | 6.07<br>(5.88-6.26) | 6.19<br>(5.87-6.51) | 5.73<br>(5.44-6.02) | 6.35<br>(5.96-6.73) |
| **Age** |  |  |  |  |  |
|  | <10y | 5.57<br>(5.32-5.83) | 5.67<br>(5.23-6.12) | 5.21<br>(4.84-5.58) | 5.96<br>(5.46-6.47) |
|  | 10-19y | 6.48<br>(6.02-6.94) | 6.33<br>(5.6-7.06) | 6.30<br>(5.63-6.98) | 7.00<br>(5.96-8.04) |
|  | 20-29y | 5.77<br>(5.28-6.26) | 5.8<br>(5.04-6.56) | 5.72<br>(4.84-6.6) | 5.80<br>(4.82-6.78) |
|  | 30-39y | 6.99<br>(6.41-7.57) | 6.84<br>(5.96-7.72) | 7.05<br>(6.00-8.09) | 7.20<br>(6.05-8.35) |
|  | 40-49y | 6.86<br>(6.08-7.65) | 7.23<br>(5.85-8.61) | 6.67<br>(5.51-7.82) | 6.47<br>(5.01-7.92) |
|  | 50-59y | 5.80<br>(4.90-6.70) | 5.76<br>(4.66-6.87) | 5.77<br>(3.95-7.59) | 5.90<br>(3.74-8.06) |
|  | 60y + | 5.84<br>(4.69-6.99) | 7.73<br>(5.54-9.91) | 3.56<br>(2.84-4.27) | 5.80<br>(4.26-7.34) |
| **Sex** |  |  |  |  |  |
|  | Male | 6.15<br>(5.87-6.43) | 6.15<br>(5.69-6.61) | 6.02<br>(5.55-6.49) | 6.34<br>(5.77-6.9) |
|  | Female | 5.99<br>(5.73-6.24) | 6.23<br>(5.79-6.67) | 5.46<br>(5.11-5.82) | 6.36<br>(5.84-6.89) |
| **Occupation** |  |  |  |  |  |
|  | Student | 6.57<br>(6.10-7.04) | 6.51<br>(5.63-7.38) | 6.24<br>(5.61-6.88) | 7.20<br>(6.25-8.15) |
|  | Office/Shop worker | 7.66<br>(6.57-8.75) | 7.75<br>(3.97-11.53) | 7.6<br>(5.93-9.27) | 7.72<br>(6.16-9.28) |
|  | Agriculture | 7.02<br>(6.35-7.68) | 7.35<br>(6.53-8.16) | 5.92<br>(4.86-6.98) | 7.00<br>(5.04-8.96) |
|  | Unemployed/Retired | 5.49<br>(5.29-5.70) | 5.58<br>(5.25-5.92) | 5.13<br>(4.81-5.45) | 5.85<br>(5.42-6.27) |
|  | Others | 5.84<br>(5.34-6.35) | 5.91<br>(5.08-6.75) | 5.95<br>(5.1-6.8) | 5.63<br>(4.63-6.63) |
| **Setting** |  |  |  |  |  |
|  | Household | 2.8<br>(2.68-2.92) | 2.94<br>(2.74-3.15) | 2.48<br>(2.28-2.67) | 3.02<br>(2.8-3.24) |
|  | Community | 3.27<br>(3.09-3.45) | 3.25<br>(2.95-3.54) | 3.25<br>(2.97-3.53) | 3.33<br>(2.98-3.67) |
| **Traveled to a different neighborhood in the month prior the interview** |  |  |  |  |  |
|  | Yes | 6.21<br>(5.83-6.59) | 6.08<br>(5.34-6.81) | 5.66<br>(5.16-6.16) | 7.22<br>(6.46-7.97) |
|  | No | 6.01<br>(5.79-6.23) | 6.22<br>(5.86-6.57) | 5.76<br>(5.4-6.13) | 5.93<br>(5.51-6.35) |
| **Contacts outside neighborhood** |  |  |  |  |  |

| (%) |  |  |  |  |
| --- | --- | --- | --- | --- |
| 0-14y | 0.67%<br>(0.4-1.04) | 0.38%<br>(0.14-0.94) | 0.19%<br>(0.03-0.77) | 1.80%<br>(1.03-3.09) |
| 15-59y | 3.98%<br>(3.23-4.89) | 4.54%<br>(3.36-6.07) | 3.3%<br>(2.19-4.9) | 3.91%<br>(2.47-6.07) |
| 60y+ | 2.74%<br>(1.28-5.53) | 1.18%<br>(0.20-4.63) | 0.00%<br>(0.00-0.00) | 10.34%<br>(4.28-21.84) |

For all sites, contacts outside school were predominantly reported between family members (46.1%), neighbors (25.2%), and other relatives outside the household (13.1%), while the remaining 15.5% of contacts occurred with friends, schoolmates outside school, or other unspecified categories. Individuals with a recent history of travel outside their neighborhood did not report an increased number of contacts, except for urban residents (t-test p=0.004). The mean number of contacts (excluding school contacts) reported by participants was lower in rural villages (5.73, 95%CI 5.44-6.02) with respect to both urban neighborhoods (6.35, 95%CI 5.96-6.73) and remote settlements (6.19, 95%CI 5.87-6.51). In particular, the mean number of daily contacts reported by the elderly (60+ years old) was much higher in remote settlements and urban neighborhoods than in rural villages (7.7 and 5.8 vs 3.6, see Table 2).

Students reported 1,372 additional contacts in schools, resulting in a mean number of 6.07 (95%CI 4.98-7.16) daily physical contacts per child (median 3, range 0-50). There were limited differences in the mean number of school contacts across geographical contexts (6.31, 95%CI 4.13-8.50 in remote settlements; 5.70, 95%CI 4.19-7.21 in rural towns; 6.54, 95%CI 4.25-8.84 in urban neighborhoods).

The analysis of contacts by age clearly shows that subjects below 30 years of age tend to interact mostly with individuals of similar age (assortative mixing). The highest contact rates were found between school aged children (10-19 years), young adults (20-39 years) and between children below 10 years and their parents (Figure 1, and Appendix). However, a marked intergenerational mixing both within households and in the community was found, especially in remote settlements.

**Figure 1.**
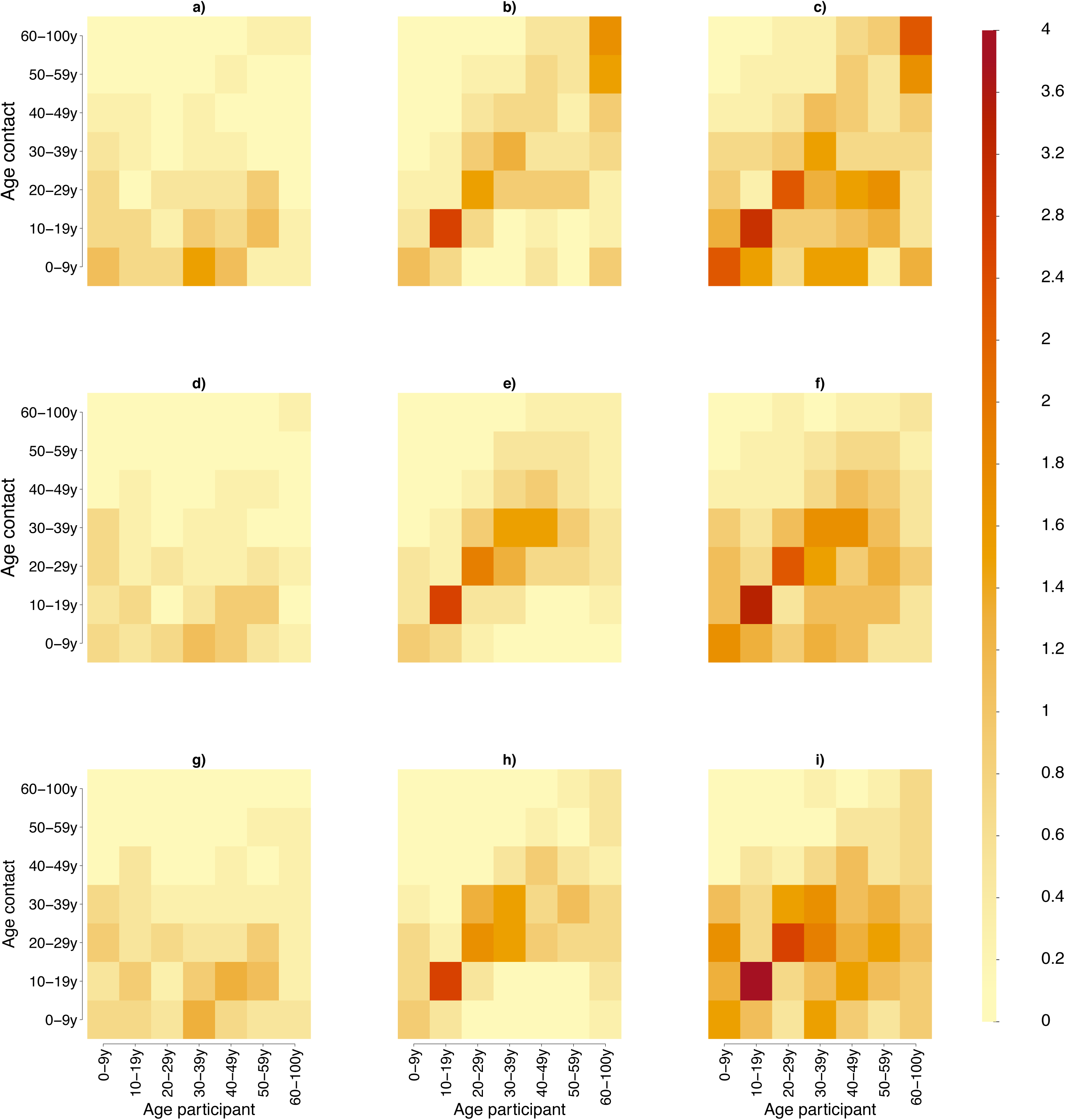
Contact matrix representing the mean number of daily contacts reported by a participant in the age group *i* with individuals in the age group *j* in household (a), in the general community (b) and both (c) in remote settlements. Panels d-f and g-I show the same quantities estimated for rural villages and for the urban neighborhoods, respectively.

## Effect of demography and age-specific contacts on COVID-19 epidemics

From the epidemic curve of reported cases, we estimated a net reproduction number *R* of 1.62 (95%CI 1.55-1.70) over approximately 6 weeks of exponential growth starting from May 1, 2020 when schools were closed in the entire country (see Appendix). We relied on this estimated of *R* to simulated COVID-19 epidemics in the SWSZ considering no school contacts. If school contacts are included, we estimate *R* to increase to 2.03 (95%CI 1.87-2.18, see Appendix), which is comparable, although slightly lower, than estimates of the basic reproduction number from other parts of the world^25,26,27,28^.

Our simulation results show that, should schools remain closed for the entire duration of the epidemic and no other interventions enacted, between 10% and 15% of the overall population would have developed respiratory symptoms or fever because of COVID-19. The fraction of critical cases (requiring mechanical ventilation and/or resulting in a fatal outcome) is estimated to be between 0.29% and 0.41% of the overall population (Figure 2). The highest prevalence of critical cases (between 4.2% and 5.4% on average) is expected within subjects aged 60 years or older. This age segment represents only about 5% of the total population in Ethiopia but is expected to represent 7 to 14% of symptomatic cases and 45% to 63% of all critical cases.

**Figure 2.**
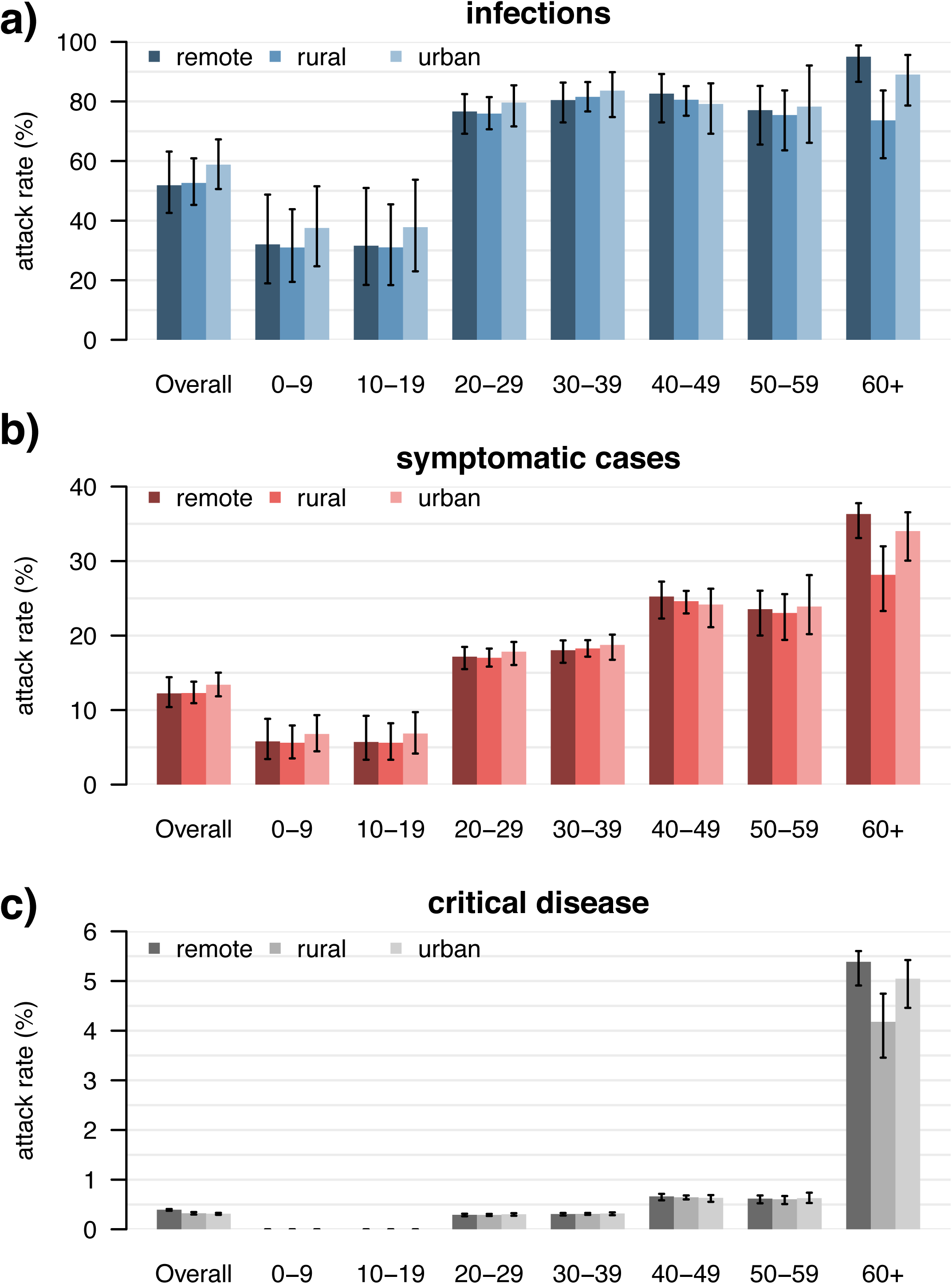
Estimated attack rates of infection (a), symptomatic cases (b), and critical disease (c), overall and by age group in different geographical contexts. We considered hypothetical epidemics with school closure and a reproduction number of 1.62 (95%CI 1.55-1.70), estimated from surveillance data^5^.

Remote settlements are expected to suffer a higher overall burden of critical cases (0.40% of the total population, 95%CI: 0.37-0.41%) compared to rural villages (0.32%, 95%CI: 0.30-0.35%) and urban neighborhoods (0.31%, 95%CI 0.29-0.33%). This difference is explained by a higher proportion of the elderly in the population (Figure 3), but also by their higher number of daily contacts and the higher intergenerational mixing (Figure 1c) compared to the other settings, which results in a higher attack rate of infections, symptomatic cases, and critical disease in this age group (Figure 2). Urban neighborhoods, where highest contact rates at younger ages were recorded, are expected to have the highest attack rate of infections (58.8%, 95%CI: 50.6-67.3) and symptomatic cases (13.4%, 95%CI: 11.8-15.0). However, since a large proportion of the overall number of infections (81.8%, 95%CI: 79.5-84.2) is concentrated on children and younger adults (up to 40 years of age), this does not result in a high overall proportion of critical disease. Finally, rural villages have lower attack rates among the elderly because of the significantly lower number of contacts reported by that age group in this geographical context (Figure 1f, Table 2).

**Figure 3.**
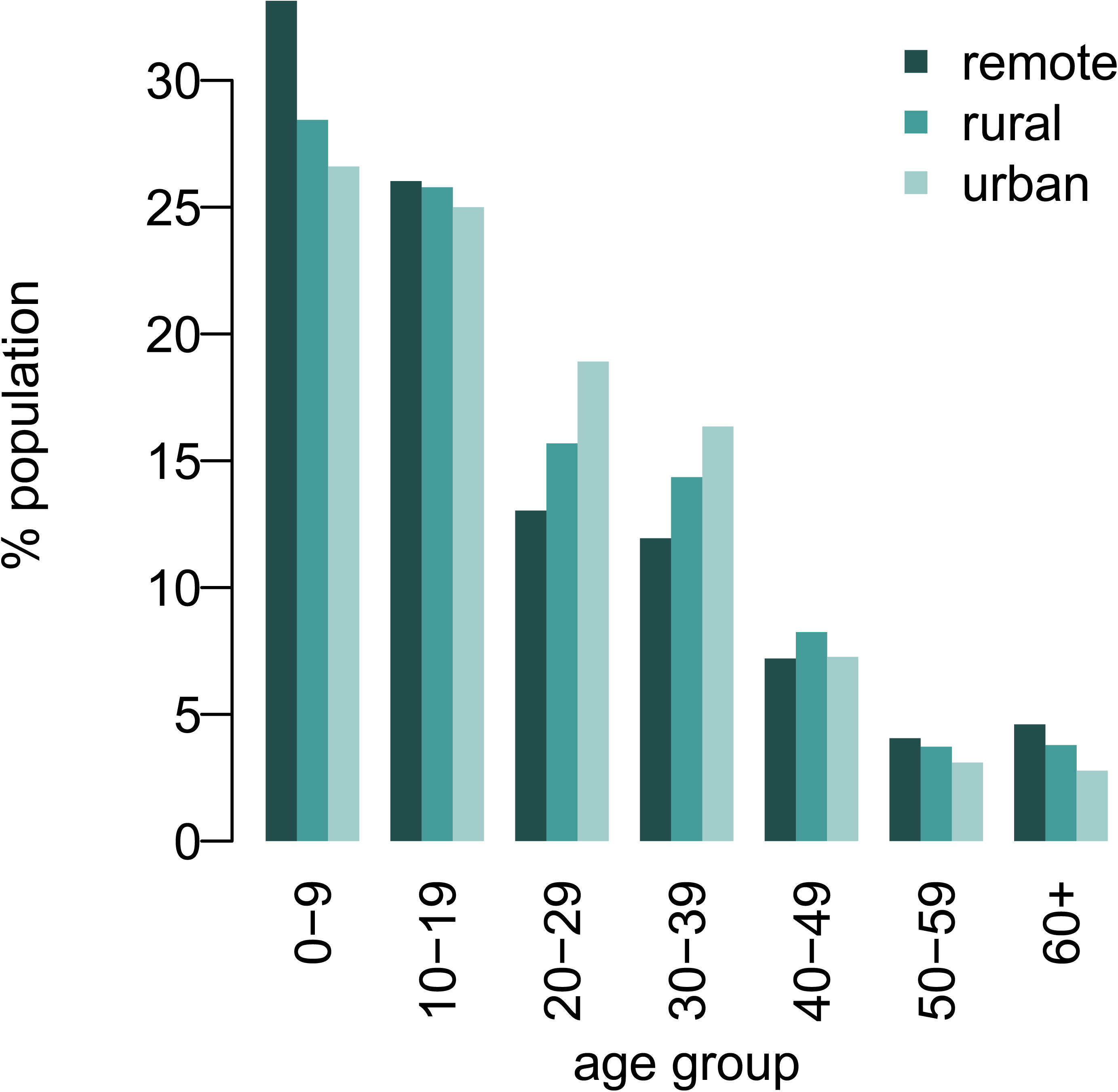
Age structure of the population in the South West Showa Zone (SWSZ), Oromia Region, Ethiopia, by geographical context, obtained from survey data.

## Discussion

Our analysis explored the effect of demographics and social contact patterns on COVID-19 burden in the South West Shewa Zone of the Oromia region, Ethiopia. Data collected within an interview-based survey highlighted differences in demographic structure and in age-specific contacts between urban neighborhoods, rural villages, and remote settlements, and were used to inform an epidemic model simulating the transmission dynamic of SARS-CoV-2. On the basis of the trajectory of COVID-19 cases observed in the country up to June 12, 2020, we estimated that between 3.1 and 4.0 patients per 1,000 inhabitants may experience critical disease (i.e., requiring mechanical ventilation and/or resulting in a fatal outcome) at the end of an epidemic mitigated by school closure alone. Considering the low availability and accessibility of healthcare, especially in remote and rural settlements, and the lack of intensive care units to treat critical patients^2,29^, it is possible that a large fraction of those cases would result in a fatal outcome, adding up to the already high background mortality rate in the region (estimated at about 6.4 per 1,000 per year^30^).

Considering the extreme scenario where all critical cases would result in a fatal outcome, we obtain an estimate of the infection-fatality ratio (IFR) ranging between 0.53% in urban neighborhoods and 0.76% in remote settlements. Such estimates are generally lower than the IFR estimated from serological studies for higher income countries^31,32^. This difference is partially due to the younger age structure of the Ethiopian population, where only 5% of individuals are older than 60 years (compared to over 20% in most of Europe^33^). However, by simply adjusting the age-specific IFR to the local demographics, Ghisolfi et al.^34^ estimated a four-fold reduction in the overall IFR in Eastern Africa with respect to European countries, which is around 2 time lower than our estimates. In fact, our simulations not only account for the demography of the population, but also for its mixing patterns.

Indeed, we found that in Ethiopia the effects of a younger population is partially compensated by high infection attack rates in the elderly, which derive from the intense intergenerational mixing and the larger number of contacts observed among the elderly. In particular, we show that these characteristics are especially marked in remote settlements, where the highest incidence of critical disease is expected to occur.

To properly interpret the results presented in our study, it is important to consider the following limitations. First, the target study population may be not representative of all Ethiopia, and in particular of epidemic patterns observed in highly urbanized areas such as the capital Addis Abeba. Second, the net reproduction number was estimated from national surveillance data^9^. This data reports cases aggregated at the country level and may suffer from a number of biases: it does not account for reporting delays; the growth over time in the number of cases may partly be ascribable to the increase in testing capacity; total cases represent the superimposition of different, asynchronous epidemics in multiple parts of the country, a majority of which coming from the highly urbanized Addis Abeba area^9^. However, we show that, assuming no restriction to school contacts, the reproduction number estimated by the model is in the range 1.87-2.18, just slightly lower than estimates of the SARS-CoV-2 basic reproduction number from other countries ^25,26,27,28^.

Moreover, our conclusions remain robust when considering a 20% increase or a 20% decrease of the reproduction number. In this case, we estimated an attack rate of critical cases ranging from 0.24 to 0.37 for rural villages and from 0.34 to 0.42 for remote settlements (see Appendix). Third, the model lacks of spatial structure. The finding from the survey that about 97% of recorded contacts have occurred within the participant’s neighborhood of residence (Table 2) suggests that local containment or confinement of COVID-19 outbreaks in rural regions of Ethiopia may be favored by low human mobility. On the other hand, the observation of a large number of cases in all regions of Ethiopia^9^ may imply that a significant widespread diffusion of the epidemic, possibly sustained by a high fraction of asymptomatic infections (Figure 2), is already ongoing. Fourth, the role played by children in the transmission of SARS-CoV-2 infections is still poorly understood and highly debated^19,35^. In the main analysis we assumed that the probability of transmission is homogeneous across all ages; however, an alternative assumption in which children are assumed half as infectious as adults would result in similar attack rates of critical cases (see Appendix S1). These results are also robust with respect to the assumption of a homogeneous susceptibility across age groups (see Appendix S1). Finally, in absence of direct data from sub-Saharan Africa, the age-specific susceptibility and proportions of infections resulting in symptomatic cases or critical disease were estimated from data from China or Europe^19,24^. However, the high prevalence of comorbidities which are uncommon in higher income countries (e.g., malnutrition^36^, tuberculosis, and malaria) and inequalities in the access to primary care represent additional vulnerabilities for African settings^2^ and may result in an underestimation of the expected disease burden. Further studies are warranted regarding the severity of COVID-19 in sub-Saharan populations, and especially in relation to its understudied potential interaction with specific comorbidities.

## Conclusions

This study provides novel data on mixing patterns in rural Ethiopia and highlights the potential impact of COVID-19 epidemics in less urbanized regions of the country. We provide estimates on the potential burden of COVID-19 under the assumption of a mitigated, but not controlled epidemic. We conclude that, although the overall mortality might be generally lower in sub-Saharan Africa compared to high income settings, thanks to younger demographics^34,37,38^, this effect may be partially offset in rural areas by higher attack rates in elderly individuals, due to high rates of intergenerational mixing. The observed contact patterns suggest that elderly individuals in remote settlements may be even more exposed to the risk of infection (and thus of critical disease), which is especially worrysome in light of the major obstacles in access to healthcare for those populations^2^.

## DECLARATIONS

### Ethics approval and consent to participate

The study was approved by the Oromia Regional Health Bureau. Written informed consent was sought for literate individuals aged 18 years or more, and from a parent or carer for younger individuals. In addition, assent was sought from all participants under 18 years of age, illiterate participants and participants who cannot read in the Oromyffa language. A written confirmation of verbal assent by a witness elected by the study participant was sought.

### Consent for publication

Not appplicable

### Availability of data and material

Data analysed during this study will be included in this published article as supplementary information files.

### Author’s contribution

FT, FM, GP, MA, SM and PP conceived and designed the study. FT, VM, WNG,AT, AG and PP collected the data. FT, MG, AZ and PP analysed the data. FT, GG, MG and AZ performed the experiments. FT, GG and PP drafted the first version of the manuscript. All authors contributed to the interpretation of the results and edited and approved the final manuscript.

## Data Availability

Data will be made available after acceptance of the manuscript on a peer-review journal

## Acknowledgments

Not applicable.

