## Supplementary material for "Demography, social contact patterns and the COVID-19 burden in different settings of Ethiopia: a modeling study": SI Appendix

### Appendix S1

#### Demography, social contact patterns and the COVID-19 burden in different settings of Ethiopia: a modeling study

Filippo Trentini[1], Giorgio Guzzetta[1], Margherita Galli[1,2], Agnese Zardini[1,3], Fabio Manenti[4], Giovanni Putoto[4], Valentina Marziano[1], Worku Nigussa Gamshie[5], Ademe Tsegaye[5], Alessandro Greblo[4], Alessia Melegaro[6,7], Marco Ajelli[8,9], Stefano Merler[1], Piero Poletti [1,*]

[1] Bruno Kessler Foundation, Trento, Italy
[2] University of Udine, Udine, Italy
[3] University of Trento, Trento, Italy
[4] Doctors with Africa CUAMM, Padova, Italy
[5] Doctors with Africa CUAMM, Ethiopia, Italy
[6] Dondena Centre for Research on Social Dynamics and Public Policy, Bocconi University, Milan, Italy
[7] Department of Social and Political Sciences, Bocconi University, Milan, Italy
[8] Department of Epidemiology and Biostatistics, Indiana University School of Public Health, Bloomington, IN, USA
[9] Laboratory for the Modeling of Biological and Socio-technical Systems, Northeastern University, Boston, MA USA

### Contents

1. Study design
2. Sample size definition
3. Data collection
4. Transmission model and reproduction numbers
5. Uncertainty in contact matrices
6. Additional results on contact patterns
7. Sensitivity analyses
8. References

### 1. Study design

The study population consisted of individuals residing in four districts (woreda) of the South West Shewa Zone (SWZS) in the Oromia region of Ethiopia. These woredas count 449,460 inhabitants and represent the main catchment area of the St. Luke Hospital located in Woliso Town. The St. Luke Hospital is a well-resourced health facility and represents the referral hospital for the entire SWSZ, serving a population of about 1.3M inhabitants with 200 beds and an annual average bed-occupation rate of 84% [1].

Data on individuals' mixing patters and local demography were collected through a cross-sectional survey, by adopting a two-stage stratified random sampling of study participants by location and age group. For each woreda, two neighbourhoods (kebeles) were identified as representative of the considered woreda, chosen as extremes illustrative socio-demographic contexts within the woreda in terms of urbanization, population density, work and travel opportunities, and distance to healthcare facilities. The target sample size was uniformly distributed across the 8 selected kebeles. The sample stratification was designed to capture different activity levels (e.g. movements, schooling/working, etc.) and the different role played by individuals in the community (e.g. household heads, women, etc.), taking into account the local schooling system (age at enrolment in pre-primary, primary, and secondary school). Individuals of all ages living in the selected sites were considered eligible for inclusion in the study. A target sample size was defined for the following age groups: <1 year old, 1-3 years old, 4-10 years old, 11-14 years old, 15-29 years old, 30-49 years old, >50 years old.

Random sampling of households and study participants was applied, using a list of predefined quotas for each site, sex and age group. Specifically, the target sample for each age group and location was equally divided into males and females. One individual per household was selected and interviewed. If the study participant was temporarily outside the household, another attempt was made later in the day or within three days from the first visit. After the second attempt, the study participant was replaced.

### 2. Sample size definition

For each age group *i*, we chose an equal sample size $n_i$ in such a way to detect, given a specified power p and significance level set at 0.05, a significant difference in the average number of contacts between at least two out the seven age groups defined above in a one-way ANOVA [2].

The optimal sample size can be computed as a function of the power of the test, the significant level and the effect size *f*, which in turn can be calculated using the following formula,

$$f = \sqrt{\frac{\sum_{i=1}^{k} \frac{1}{k}(\mu_i - \mu)^2}{\sigma^2}},$$

where *k* is the number of groups, $\mu_i$ is the expected average number of contacts for age group *i*, $\mu$ is the expected average number of contacts in the overall population, and $\sigma^2$ is the expected constant error variance within groups. As shown in the Figure 1, setting a power of 80%, k=7 and a significance level at 0.05, a sample size of 120 in each group would correspond to the optimal sample size for *f*=0.13, which can be considered as a sufficiently small effect size. Indeed, by considering values of $\mu_i, \mu$ and $\sigma^2$ obtained in previous studies on social contacts [3,4], the effect size would be around 0.17. Based on previous findings available at the time [3], the considered sample size enabled the detection of 20% difference in the average number of daily contacts by age group.

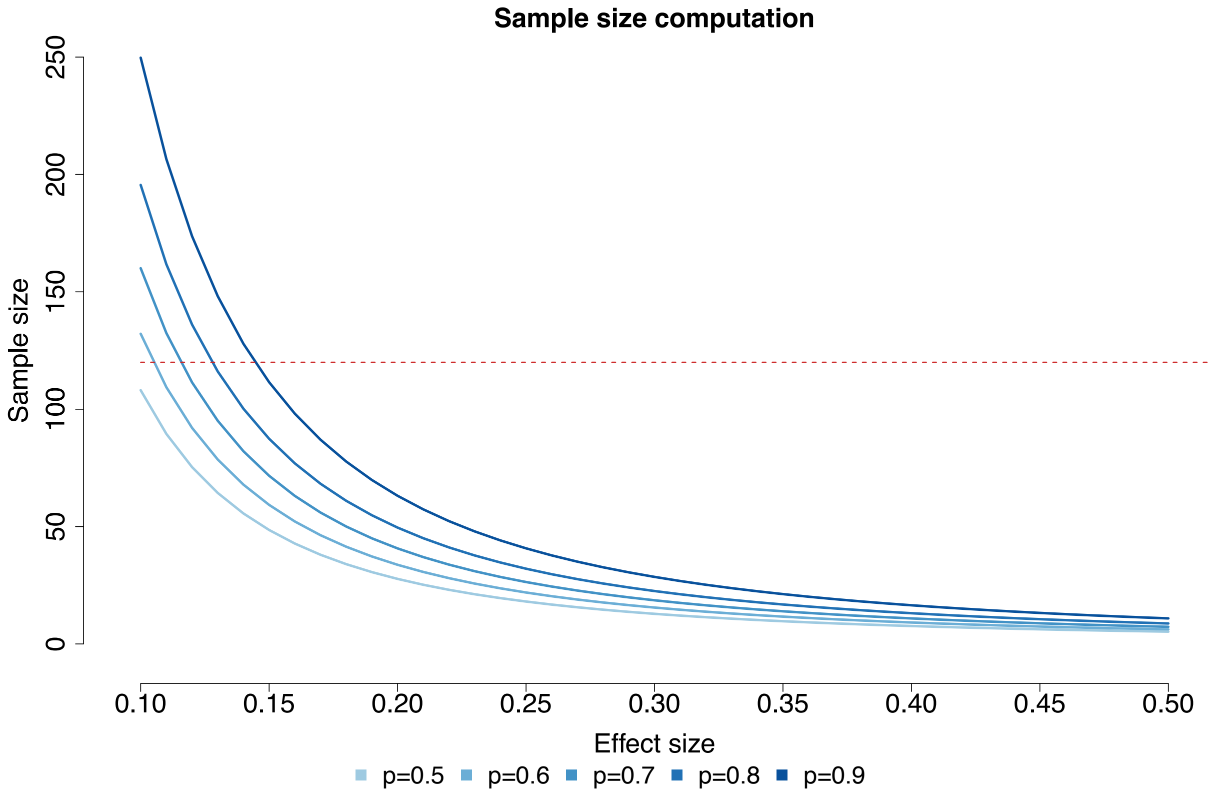


**Figure S1.** Optimal sample size computed for different values of effect size and power of the test (p), assuming a significance level of 0.05. The horizontal line represents the target sample size defined in our study.

In the final sampling scheme, senior adults (30-49 years old) were slightly over-sampled and the elderly (>50 years old) slightly under-sampled, due to the different relative frequency in the population.

#### 3. Data collection

For each study participant we collected data on their age, sex, household size, household composition, place of residence and the full list of contacts they experienced in the day preceding their interview. Specifically, the frequency and type (either physical or non-physical) of each social encounter was collected, along with the age and relationship with each listed contact and the transmission setting and the kebele (neighborhood) where the interaction occurred. The day of the week in which the interview was administered to each study participant was also recorded.

Data collection was performed through interviews of the study participants by field investigators, who directly inserted anonymized answers into an electronic dataset based on Survey CTO software, installed on tablets. Questions were designed in English and translated into the predominant local language (Oromyffa). Data entry conflicts and inconsistencies were identified automatically by the system and resolved as the data entry progressed.

Results of a preliminary pilot study on a sample of 20 people, recruited in a different site from those used in the survey, were used to optimize the interview, address logistic challenges and refine the operational guidelines for the data collectors conducting the interviews. Following the results obtained for students in the pilot, we decided to collect only data on physical contacts at school. In particular, only the overall number of contacts experienced by students at school was collected. Responses gathered during the pilot were excluded from the analysis. Quality of data collection was then verified by administering 78 individual interviews in a remote settlement of the SWSZ which was outside the original target sites. These data were included in the analysis of contact patterns.

#### 4. Transmission model and reproduction numbers

We developed a transmission model for the spread of SARS-CoV-2 infection, based on an age-structured susceptible-infectious-removed (SIR) scheme. Contact data collected with 938 individual interviews was used to inform the model with the age-specific mixing patterns in the South West Shewa Zone across different geographical contexts and transmission settings. To such an aim, participants and contacts were grouped in six 10-year age classes plus an additional class including all individuals aged 60 years or older. When exact age of the contactee was unknown, the midpoint of the age range provided during the interview was used to assign the contactee to an age class. The age of contacts experienced by participants at school was inferred using information on the age of students attending their school and grade. We then computed age-specific contact matrices $C^x_{a,\tilde{a}}$ representing the average number of contacts reported by respondents in age group $a$ with contactees in age group $\tilde{a}$ in the setting $x$. Considered transmission settings included the household, the school and the general community. Contacts at work were aggregated with all other contacts occurring in the community, since for people employed in agriculture (about 33% in Ethiopia) and many other occupations (e.g. street vendors and people participating to community markets) it was difficult to disentangle encounters occurred because of their job from other random contacts. Only physical contacts were considered for school; both physical and non-physical social interactions were considered for other transmission settings. Sample variability was explored using bootstrap sampling, as detailed in the following section. Contact matrices were separately computed for three different geographical contexts by aggregating interviews conducted in remote settlements (n=400), rural villages (n=326) and the two urbanized neighbors of Woliso Town (n=212).

In the model, infectious contacts within and between age classes may occur in three different transmission settings (home $H$, schools $S$, community $C$), and are combined in an overall contact matrix according to the following equation:

$$\text{[Eq1]} \quad M_{a,\tilde{a}}(t) = C^H_{a,\tilde{a}} + \delta_s\, C^S_{a,\tilde{a}} + C^C_{a,\tilde{a}}$$

where:

- $C^H_{a,\tilde{a}}, C^S_{a,\tilde{a}}, C^C_{a,\tilde{a}}$ are the contact matrices for the transmission settings described above;
- $\delta_s$ is a parameter which is set equal to 0 to consider the transmission dynamics under the school closure mandate, and equal to 1 to assess the transmission potential when schools are open;
- $M_{a,\tilde{a}}(t)$ represents the age-specific contact matrix, whose entries describe the mean number of persons in age group $\tilde{a}$ encountered by an individual of age group $a$ per day across different settings.

The proportions of the SWSZ population living in each geographical context were used as sampling weights to compute average contact matrices for the entire SWSZ.

In the model, we assumed asymptomatic and symptomatic individuals to be equally infectious, as suggested by an early analysis of virological data from Lombardy [5] and Veneto [6]. The transmission model considers three consecutive infectious compartments to reproduce a gamma-distributed generation time [5,7]. The force of infection for subjects of age $a$ is defined as:

$$\text{[Eq2]} \quad \lambda_a(t) = \beta r_a \sum_{\tilde{a}} \check{r}_{\tilde{a}} M_{a,\tilde{a}} \frac{\alpha_I I_{\tilde{a}}(t) + \alpha_J J_{\tilde{a}}(t) + \alpha_K K_{\tilde{a}}(t)}{N_{\tilde{a}}}$$

where:

- $\beta$ is a scaling factor shaping the number of potentially infectious contacts resulting in infection;
- $r_a$ is the relative susceptibility to SARS-CoV-2 infection at age $a$;
- $\check{r}_{\tilde{a}}$ is the relative infectiousness at age $\tilde{a}$;
- $I_{\tilde{a}}(t)$, $J_{\tilde{a}}(t)$ and $K_{\tilde{a}}(t)$ represent the number of individuals of age $\tilde{a}$ in the three stages of infection I, J, K, at time t.
- $\alpha_I, \alpha_J$ and $\alpha_K$ are adjusting factors for individuals' infectiousness during the three stages of infection I,J and K;
- $N_{\tilde{a}}$ represents the total number of individuals in age group $\tilde{a}$.

In the baseline analysis, we assumed that, compared to adults aged 20-59 years ($r_a = 1$), individuals aged <20 years are 67% less susceptible to infection (i.e. $r_a = 0.33$; 95%CI 0.24-0.47) and those aged ≥60 years are 47% more susceptible to infection ($r_a = 1.47$; 95%CI 1.16-2.06) [8]; homogeneous susceptibility to SARS-CoV-2 infection across ages was considered for sensitivity analysis ($r_a = 1$ for all $a$). In the baseline analysis, individuals of different ages were considered equally infectious ($\check{r}_{\tilde{a}} = 1$ for all $\tilde{a}$). For sensitivity analysis, we assume that individuals aged 0-19y are 50% less infectious than other individuals ($\check{r}_{\tilde{a}} = 0.5$ when $\tilde{a} < 19$; $\check{r}_{\tilde{a}} = 1$ for $a \geq 20$). Finally, we assumed that recovering from infection provides full immunity against re-infection for at least the duration of our simulations (2 years).

Transitions across different epidemiological classes can be summarized by the following differential system:

$$[\text{Eq3}] \quad \begin{cases} S_a'(t) = -\lambda_a(t)\, S_a(t) \\ I_a'(t) = \lambda_a(t) S_a(t) - \gamma I_a(t) \\ J_a'(t) = \gamma I_a(t) - \gamma J_a(t) \\ K_a'(t) = \gamma J_a(t) - \gamma K_a(t) \\ R_a'(t) = \gamma K_a(t) \end{cases}$$

where:

- S represents the number of individuals susceptible to SARS-CoV-2 infection;
- $\gamma$ is the recovery rate associated with each stage of infection: I, J, K;
- R represents the number of individuals who recover from the infection.

We assumed that the average generation time of SARS-CoV-2 can be approximated with the observed average serial interval, which was estimated in 6.6 days [5]. As the average generation time in a model with three stages of infectiousness is given by $2/\gamma$ [9], we assumed $\gamma$=0.303 days$^{-1}$. The adjusting factors $\alpha_I, \alpha_J$ and $\alpha_K$ were set at 0.014, 0.9 and 0.086, respectively to reproduce a distribution of the generation time consistent with that of the observed serial interval, i.e. a Gamma distribution with shape 1.87 and rate 0.28 [5,9]. A comparison between the model generation time and the observed SARS-CoV-2 serial interval is shown in Figure S2.

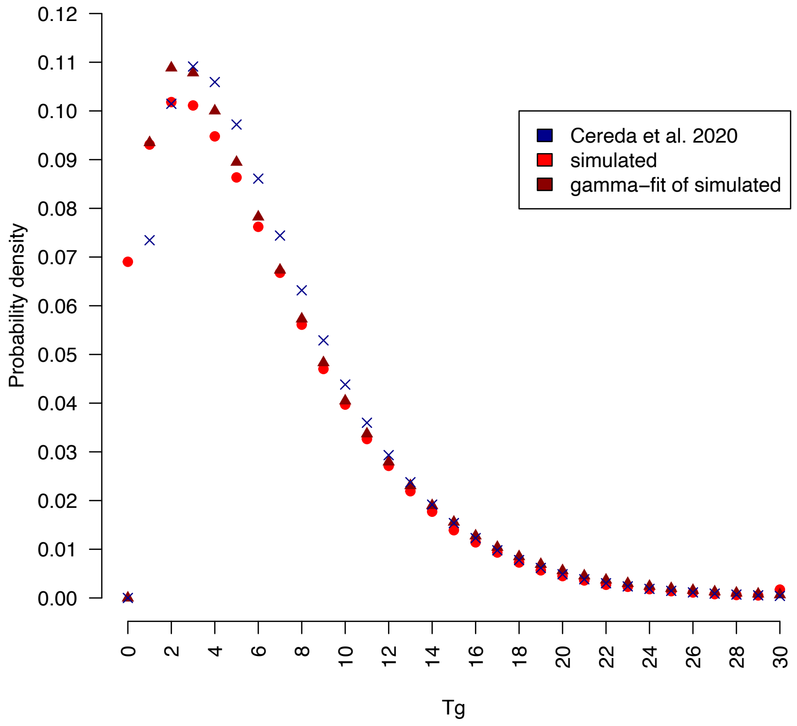

**Figure S2.** Distribution of the SARS-CoV-2 generation time (red) as simulated in our model when assuming $\gamma$=0.303 days$^{-1}$, $\alpha_I = 0.014, \alpha_J = 0.9$ and $\alpha_K = 0.086$ compared to the distribution of the SARS-CoV-2 serial interval as observed in Italy (blue).

Reproduction numbers (R) associated with different geographical contexts were computed by using the Next Generation Matrix approach [10]. The parameter $\beta$ was assumed to be equal across different geographical contexts and calibrated by considering the average contact matrix for the entire South West Shewa Zone, by computing the model's Next Generation Matrix under the assumption of school closure, and by assuming that the resulting R is equal to the reproduction number estimated from the initial (from May 1 to June 12) exponential growth characterizing the reported COVID-19 cases in Ethiopia (mean 1.62; 95%CI: 1.55-1.70, see Figure S3) [11]. Alternative values of R (i.e +/- 20%) were considered for sensitivity.

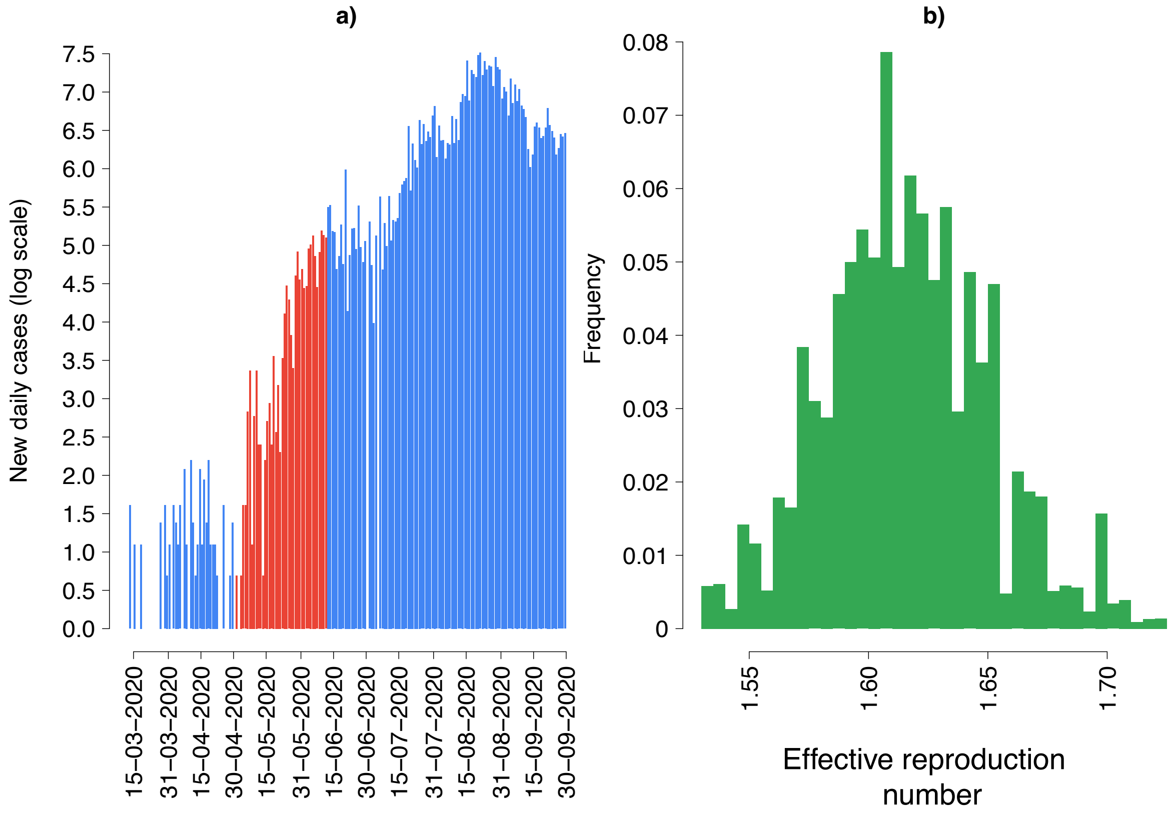


**Figure S3.** a) Daily COVID-19 cases reported in Ethiopia [12]. The red bars show the exponential phase considered to estimate the SARS-CoV-2 reproduction number in Ethiopia. b) Estimates of R obtained from the exponential growth of cases observed between May 1 and June 12.

Dynamic transmission of SARS-CoV-2 was investigated separately for the three geographical contexts (remote settlements, rural villages and urban neighbors) by considering a population stratified into 7 age groups (six 10-year age groups from 0 to 59 years and one age group for individuals aged 60 years or older). The age distribution of household members of study participants was used to define the population age-structure across different geographical contexts.

Simulation results shown in the main text and in the following sections were obtained by using a stochastic version of the model described above and 1,000 stochastic runs accounting for variability in available estimates of $r_a$ [8], uncertainty in the derived contact matrices and the uncertainty in the estimated value of R from surveillance data (Figure S3). Each simulation was initialized with 5 infections every 10,000 inhabitants, assigned randomly across age classes.

Age specific attack rates for symptomatic infections and critical cases were obtained by applying estimates for the absolute probability of developing symptoms (respiratory or fever), and critical disease (either requiring mechanical ventilation or resulting in death) after infection, as provided in [13].

#### 5. Uncertainty in contact matrices

In order to take into account sample variability, we computed 1,000 bootstrapped contact matrices for each geographical contexts and transmission setting. At each bootstrap iteration, we sampled with replacement 400, 326 and 212 interviews from those obtained in remote settlements, rural villages and urban neighborhoods respectively, choosing the age of the participant with probability proportional to the age distribution of the Ethiopian population [14]. Then, we counted for each participant *i* of age group $a$ the number of contacts reported with individuals of age $\tilde{a}$ in the setting $x$, $c^x_{a,\tilde{a}}(i)$, and estimated the average number of contacts occurring in the setting $x$ between ages $a$ and $\tilde{a}$ from the following equation:

$$[\text{Eq4}]\ C^x_{a,\tilde{a}} = \frac{\sum_{i=1}^{P_a} c^x_{a,\tilde{a}}(i)}{P_a}$$

where $P_a$ is the number of sampled participants of age group $a$.
Contact matrices resulting by averaging entries of 1,000 bootstrap $C^x_{a,\tilde{a}}$ are reported in Figure S4 and Figure S5.

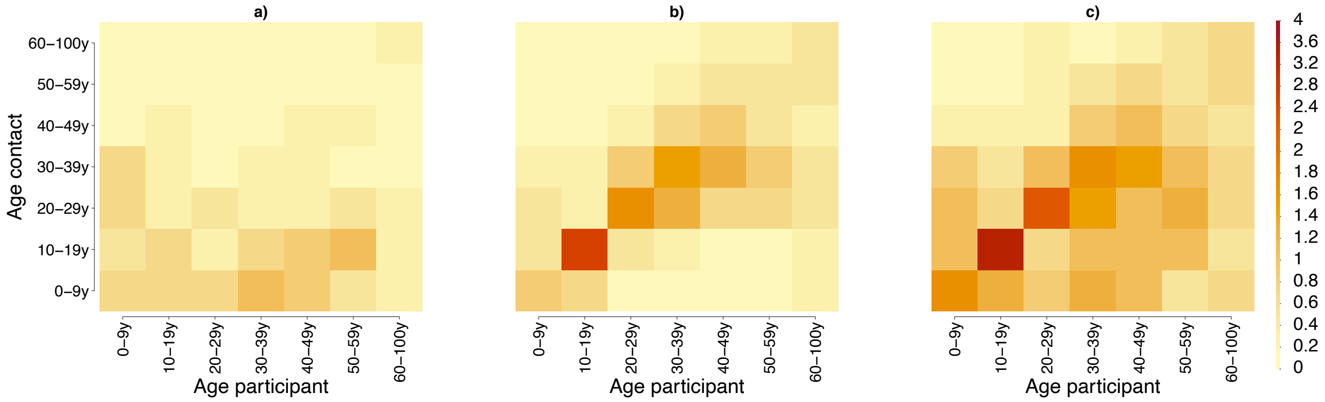


**Figure S4.** Age-specific contact matrices as obtained by averaging 1,000 bootstrapped contact matrices representing the average number of daily contacts reported by participants in the age group *i* with individuals in the age group *j* in household (a), in the general community (b) and both (c) in the SWSZ.

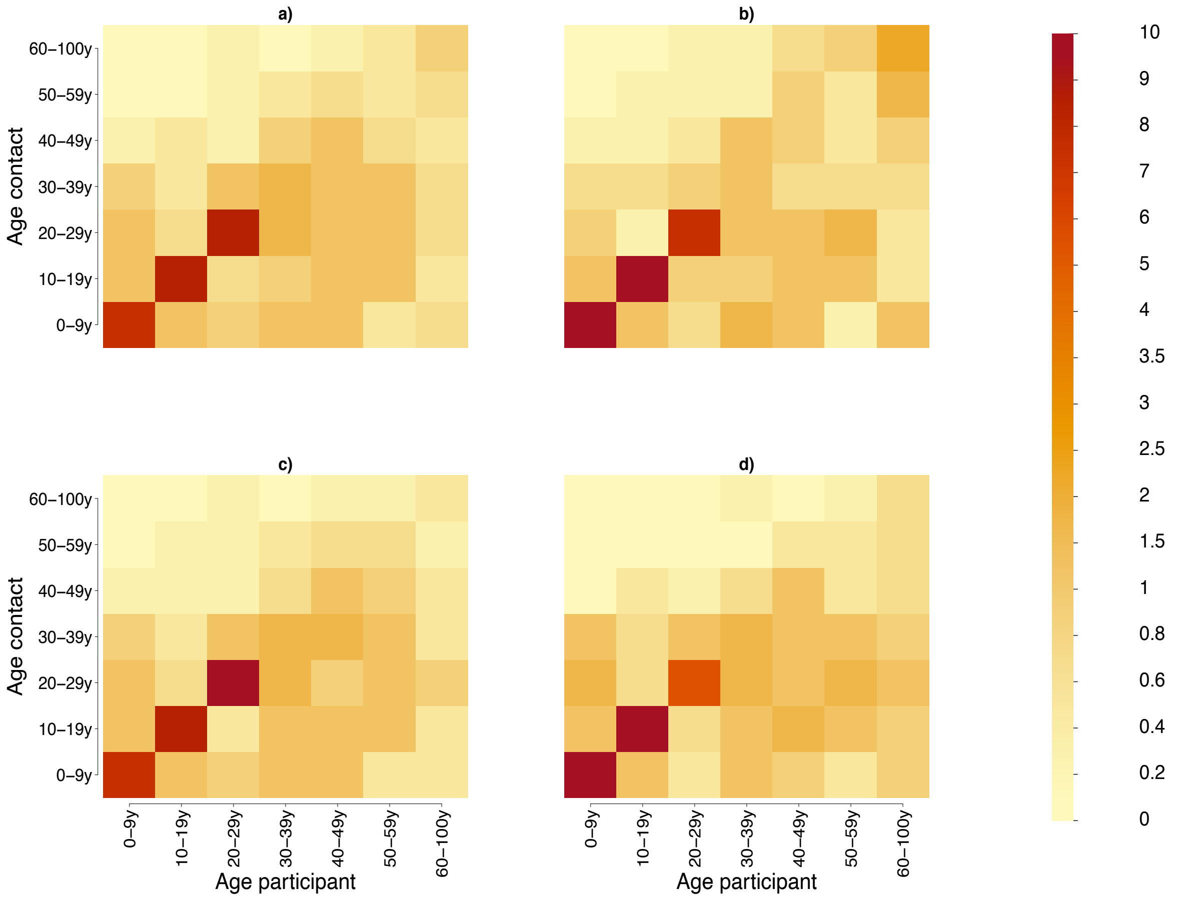


**Figure S5.** Age-specific contact matrices as obtained by averaging 1,000 bootstrapped contact matrices, representing the average number of daily contacts reported by participants in the age group *i* with individuals in the age group *j* across all settings (including schools) in the entire SWSZ (a) in remote settlements (b), rural villages (c) and urban sites (d).

#### 6. Additional results on contact patterns

The mean number of daily contacts per person was analyzed with respect to a set of covariates, including age, sex, type of work and geographical context of the study participant, and day of the week in which the encounter occurred. A statistical comparison of mean values was carried out using either t-tests or ANOVA if the strata are more than two. Differences among three or more group means were assessed by a post-hoc analysis based on the Tukey test. A Kolmogorov–Smirnov (KS) test was used to compare distributions across different strata.

In our sample, 51% of individuals were female, and no significant differences were found in the sample age distribution across different geographical contexts (pairwise KS test p-value>0.28). Differences in the age distribution of all household members of study participants are reported in Figure S6.

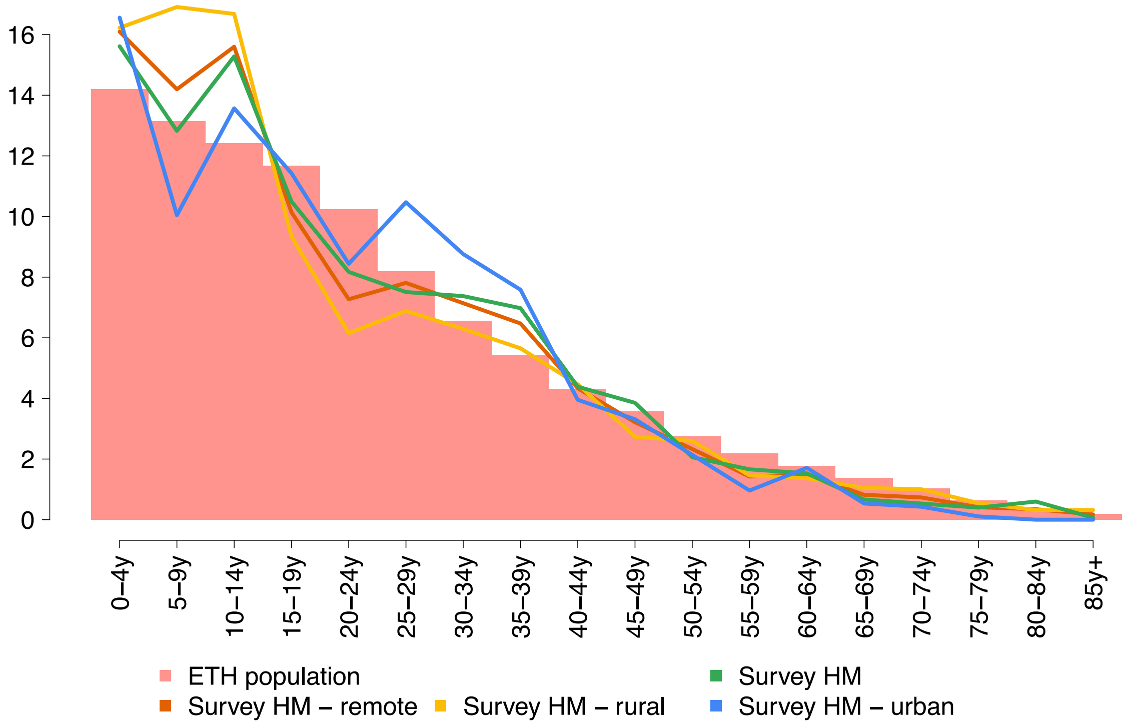


**Figure S6.** Age distribution of household members (HM) of study participants residing in the three geographical contexts and overall with respect to the age distribution of the Ethiopian population reported in [14].

Among study participants residing in remote settlements, 88.5% of male adults reported to work in agriculture. Although agriculture remains the main occupation in rural villages (30.6%), 38.8% of male adults living in these sites reported to be office, shop or manual worker; 30% of adults living in urban neighbourhoods were unemployed. In all sites, more than 60% of adult females were housewives and only 5% of working adults reported travels to a different kebele to reach their workplace. Only 9.0% of study participants accessed a health facility in the month preceding their interview. This latter percentage varies largely across age and geographical context, with percentages ranging from 7.7% among children living in remote settlements to 22.3% among those living in urban neighbors, respectively. 87.3% of the participants reported they were never admitted to the local hospital.

Highest contact rates were recorded among individuals aged 35-44 (7.22 95%CI 6.51-7.93), lowest in younger children (5.16 95%CI 4.87-5.45). However, the average number of daily contacts reported by individuals aged 65 years or more was similar to those reported by individuals aged 25-34 years (6.41, p-value>0.99). The number of daily contacts reported by people employed in agriculture was also remarkably high (mean: 7.02) when compared to office workers (mean: 8.07) and retired individuals (mean: 4.67). A similar number of daily contacts was found in males and females (6.15 vs 5.99, p-value=0.40). The number of contacts experienced during the weekends were not significantly different from those experienced during the week (6.12 vs 6.05, p-value=0.74). A significantly larger proportion of contacts outside the household was found among study participants living in rural villages (56.8%) than in those living in remote settlements (52.5% p-value =0.013) or in the more urbanized neighborhoods (52.4%, p-value =0.035). The percentage of contacts occurring outside the kebele of residence was very low in all sites: 1.5% in rural towns, 2.1% in remote settlements and 2.9% in urban neighborhoods. However, adult males residing in the urban neighborhoods and the rural towns (representing the 10.7% of the sample) were twice more likely to travel outside of their neighborhood compared to those living in remote settlements (p-value <0.001).

#### 7. Sensitivity analyses

We conducted a set of sensitivity analyses to evaluate how estimates of the potential COVID-19 burden change across different geographical contexts when assuming that:

- susceptibility to infection is homogeneous across all different age classes (i.e., assuming $r_a = 1$ for any $a$ in Eq. 2);
- the infectiousness of individuals aged between 0 and 19 years is 50% lower compared to older individuals (i.e., assuming $\check{r}_{\tilde{a}} = 0.5$ when $\tilde{a} < 20$ and $\check{r}_{\tilde{a}} = 1$ for $\tilde{a} \geq 20$ in Eq. 2) and that susceptibility to infection is heterogeneous by age, as defined for the baseline analysis;
- the reproduction number in the SWSZ is increased and decreased by 20% with respect to the value used in the baseline analysis, while susceptibility to infection and infectiousness is the same as in the baseline analysis.

Figure S7 shows the estimated attack rates of infection, symptomatic cases, and critical disease in a hypothetical epidemic with school closure, by assuming that the reproduction number in the entire SWSZ is 1.62 (95%CI 1.55-1.70), as estimated from surveillance data [12], and under the hypothesis of homogeneous susceptibility by age. Figure S8 shows the same quantities under the hypothesis that the infectiousness of individuals younger than 20 years of age is half of all other individuals.

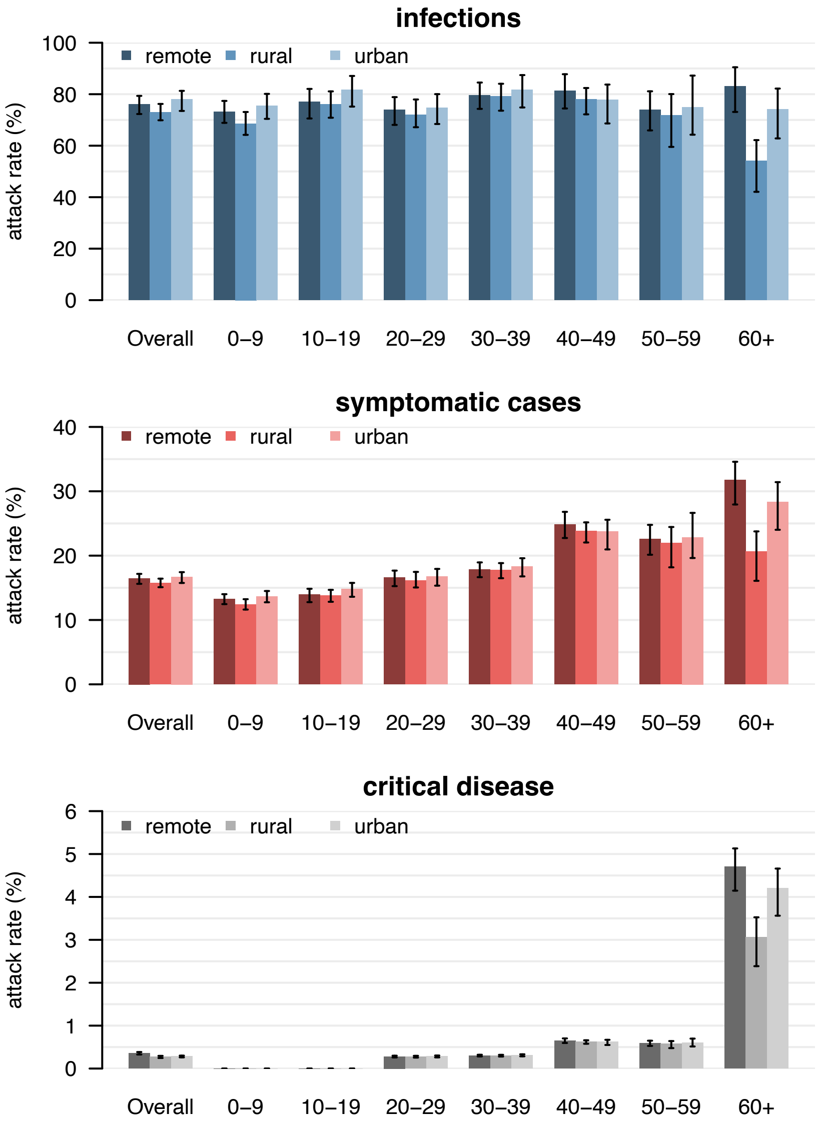


**Figure S7.** Estimated attack rates of infection (top), symptomatic cases (middle), and critical disease (bottom), overall and by age group in different geographical contexts. We considered hypothetical epidemics with school closure under the assumption of homogeneous susceptibility and that the reproduction number in the entire SWSZ is 1.62 (95%CI 1.55-1.70).

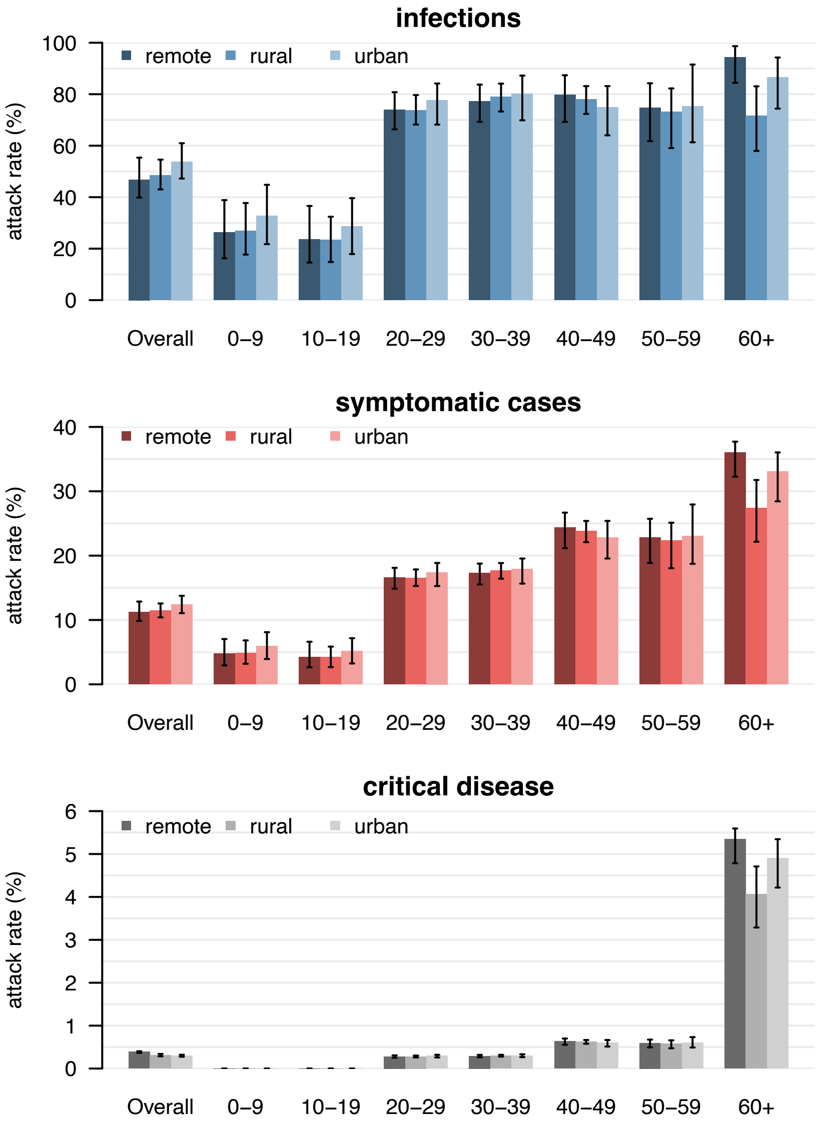


**Figure S8.** Estimated attack rates of infection (top), symptomatic cases (middle), and critical disease (bottom), overall and by age group in different geographical contexts. We considered hypothetical epidemics with school closure, under the hypothesis that the infectiousness of individuals younger than 20 years of age is half of all other individuals and that the reproduction number in the entire SWSZ is 1.62 (95%CI 1.55-1.70).

Figures S9 and S10 show the estimated attack rates of infection, symptomatic cases, and critical disease in a hypothetical epidemic with school closure when the average reproduction number in the entire SWSZ is decreased or increased by 20%.

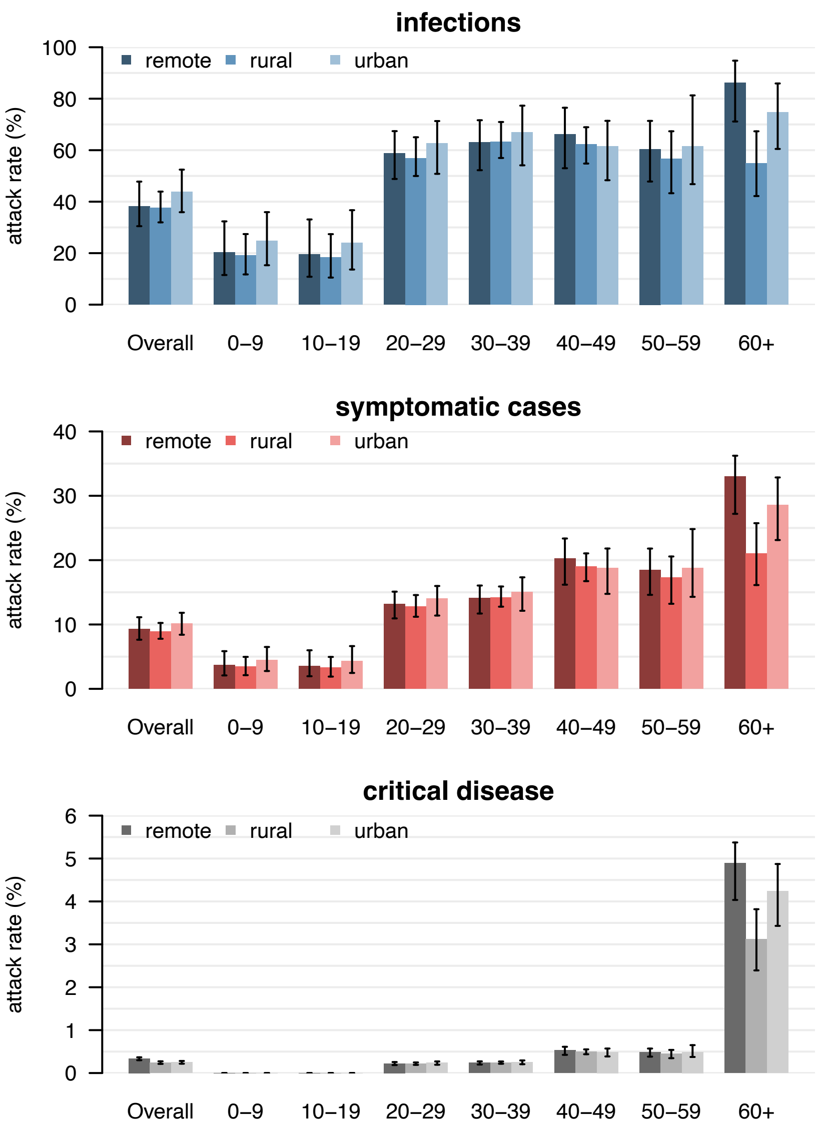


Figure S9. Estimated attack rates of infection (top), symptomatic cases (middle), and critical disease (bottom), overall and by age group in different geographical contexts when assuming a 20% decrease of the reproduction number.

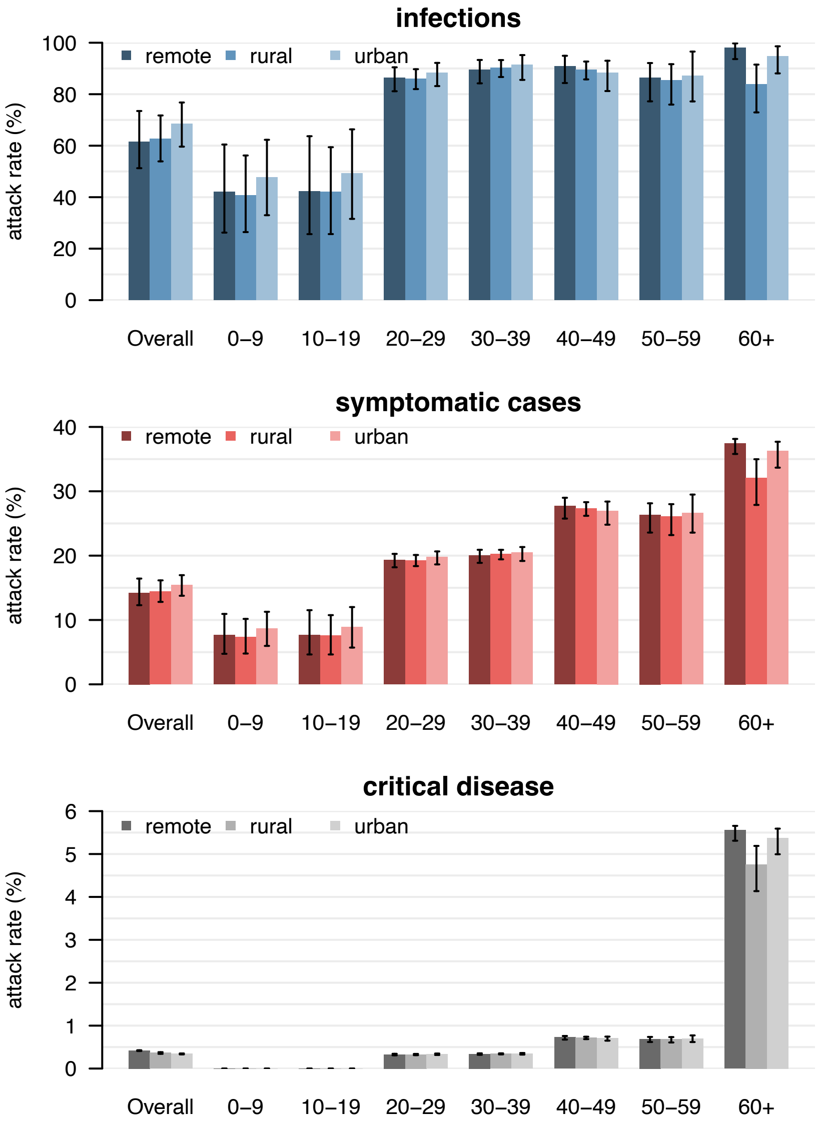


**Figure S6.** Estimated attack rates of infection (top), symptomatic cases (middle), and critical disease (bottom), overall and by age group in different geographical contexts, when assuming a 20% increase of the reproduction number.
